# Progress, stasis, and regression through the hypertension care continuum: Longitudinal evidence from population-based cohort data in four populous middle-income countries

**DOI:** 10.1101/2021.04.30.21256391

**Authors:** Nicole Mauer, Pascal Geldsetzer, Jennifer Manne-Goehler, Justine Davies, Andrew C. Stokes, Margaret McConnell, Mohammed K. Ali, Volker Winkler, Nikkil Sudharsanan

## Abstract

**Background:** Controlling and managing hypertension is a highly dynamic process yet, to our knowledge, existing evidence on hypertension control gaps in middle-income countries (MICs) is largely based on cross-sectional data. We provide the first longitudinal investigation of how individuals with hypertension move through the care continuum over time across multiple MICs.

**Methods:** We used multiple waves of population-based longitudinal cohort data from China, Indonesia, Mexico and South Africa. Based on measured blood pressure and information on hypertension diagnosis and treatment status, we classified adults aged 40 + into four care stages at both the baseline and follow-up waves: undiagnosed; diagnosed and untreated; diagnosed, treated, but uncontrolled (systolic blood pressure (SBP) ≥ 140 mmHg or diastolic blood pressure (DBP) ≥ 90 mmHg); diagnosed, treated, and controlled (SBP < 140 mmHg and DBP < 90 mmHg). We estimated the probability of individuals progressing forward or regressing backwards through the continuum over a five-to nine-year between-wave period and investigated how these probabilities varied by age, sex, household location and educational attainment using Poisson regression models. We also estimated the probabilities of important clinical transitions (e.g. becoming diagnosed or treated; achieving blood pressure control; discontinuing treatment and losing blood pressure control).

**Findings:** Our data included 8359 individuals with hypertension (China: N=1371, Indonesia: N= 3438, Mexico: N=1946, South Africa: N=1604). Across all countries, there was a less than 50% probability of forward progression through the care continuum over time. Just over one in four undiagnosed individuals became diagnosed (China 30% [95% CI 26-33%], Indonesia 30% [95% CI 28-32%], Mexico 27% [95% CI 25-29%], South Africa 37% [95% CI 34-39%]) and one in three diagnosed, untreated individuals became treated (Indonesia 17% [95% CI 14-21%], Mexico 30% [95% CI 28-32%], China 48% [95% CI 39-56%], South Africa 42% [95% CI 40-44%]). Importantly, there were very high probabilities of regressing to less advanced continuum stages: up to nine in ten treated and controlled individuals lost blood pressure control (Indonesia 92% [95% CI 88-95%], Mexico 77% [95% CI 72-81%], China 76% [95% CI 68-83%], South Africa 48% [95% CI 42-54%]) and up to three in four individuals discontinued treatment over the follow-up period (China 36% [95% CI 32-41%], Indonesia 70% [95% CI 67-73%], Mexico 34% [5% CI 32-36%], South Africa 25% [23-27%]). Individuals from rural households were disadvantaged in all countries but China, while females were more likely to progress through the continuum in Indonesia and Mexico.

**Interpretation:** Our results uncover critical gaps in hypertension care in MICs in both early and late stages of the continuum. Adopting a longitudinal perspective reveals that policies solely aimed at improving diagnosis or initiating treatment may not lead to large improvements in control, as treatment initiation rates are low and achievement of blood pressure control and adherence to therapy are rarely sustained over time.

**RESEARCH IN CONTEXT:** *Evidence before this study:* We searched PubMed for articles published from database inception until January 1st, 2021 using variations of the search terms “blood pressure”, “hypertension”, “continuum”, “cascade”, “treatment”, “diagnosis”, “control”, “treated”, “diagnosed”, “controlled” to screen titles and abstracts. The currently largest studies estimating nationally representative levels of hypertension diagnosis, treatment and control across multiple low- and middle-income countries (LMICs) are the Prospective Urban Rural Epidemiology (PURE) study from 2013, which included 140 000 individuals across 14 LMICs and a more recent study based on 1·1 million adults in 44 LMICs from 2019. Both studies highlight important gaps in awareness, diagnosis, treatment and control of hypertension across populations in LMICs. However, these and smaller non-nationally representative or single country studies are all based on cross-sectional data and none capture the dynamic nature of chronic disease care and how individuals move through the hypertension care continuum over time.

*Added value of this study:* To our knowledge, we provide the first longitudinal evidence on how individuals with hypertension in middle-income countries (MICs) move through the hypertension care continuum over time using country-wide, longitudinal cohort data from four MICs, which span three different continents and account for close to one-fourth of the world population. The longitudinal perspective provides new insights over existing cross-sectional cascades by capturing critical dynamic elements of chronic disease management, such as how individuals arrived at a specific continuum stage or whether they move forward or backward through continuum stages over time.

*Implications of all available evidence:* Our results reveal that individuals rarely sustain blood pressure control and that they tend to discontinue treatment over time. These results can inform efforts to improve hypertension control by revealing the need to move beyond policies aimed solely at screening and diagnosis to those that also aim to help individuals sustain blood pressure control over time.

## INTRODUCTION

Hypertension affects one in four adults and is the leading modifiable cause of cardiovascular disease and premature mortality worldwide.^1^ Low- and middle-income countries (LMICs) face a disproportionately high burden of hypertension: two-thirds of individuals with hypertension live in LMICs^1,2,3^ and this number is expected to increase substantially by 2050.^4^ Middle-income countries (MICs) in particular are expected to undergo considerable population ageing over the coming decades^5^, which is likely to generate a surge in people requiring hypertension care.^4^ Treating hypertension substantially reduces cardiovascular disease mortality, yet many countries have crucial gaps in hypertension diagnosis and control and only 21 MICs (out of 106 classified by the World Bank^6^) are currently on track to achieve Sustainable Development Goal target 3·4 of reducing premature mortality from non-communicable diseases (NCDs) by one third by 2030.^7,8^

Developing targeted, evidence-informed policies to manage hypertension first requires identifying the greatest bottlenecks and gaps in care. Several recent studies have used the hypertension care continuum -- the series of sequential steps from diagnosis to treated and controlled blood pressure -- as one of the primary frameworks for assessing gaps in hypertension management.^8,9,10^ The major limitation of these studies is that they are based on cross-sectional data and thus fail to capture critical dynamic elements of chronic disease management, such as how individuals arrived at a specific continuum stage or whether they move forward or backward from that stage over time. For example, individuals who are diagnosed but untreated could either have failed to progress through the continuum by not initiating treatment, or, they could have moved backwards through the continuum by stopping treatment after having initiated it previously. Identifying which pathways are more pronounced is critical for formulating effective policy since the reasons for a failure to initiate treatment are likely different from the reasons for discontinuing treatment. Relatedly, cross-sectional continuums give the impression that individuals with treated and controlled blood pressure (BP) require no additional attention and that policies should focus on progressing individuals from other continuum stages. However, reaching a state of treated and controlled BP is also not permanent, and individuals may fail to maintain blood pressure control over time due to physiological changes from aging^11^, shifting lifestyle patterns, or because they are inconsistent with or stop BP treatment. Unfortunately, to date there are few detailed longitudinal studies describing individuals’ movement through the hypertension continuum over time in MICs.

Here, we overcome this important limitation by providing, to our knowledge, the first longitudinal assessment of the hypertension care continuum in multiple MICs. We focus on four MICs (China, Indonesia, Mexico, and South Africa) with reported adult hypertension prevalence rates between 19% to 24%^2^, which span three different continents and make up close to one-fourth of the world’s population. For each country, we describe how individuals move between each of the care continuum stages over an approximately seven-year period. We also investigate if and how the transitions vary across age, sex, education, and urbanicity. Importantly, we examine both progression through the care continuum and the probability of regressing from more to less advanced continuum stages.

## MATERIALS AND METHODS

### Data

We used longitudinal cohort data from country-wide household surveys in four middle-income countries: The China Health and Nutrition Survey (CHNS), the Indonesian Family Life Survey (IFLS), the Mexican Family Life Survey (MxFLS), and the South African National Income Dynamics Study (NIDS). These surveys aim to monitor a range of population trends over time with a focus on health, social and economic wellbeing, living conditions, and demographic changes. Importantly, all four surveys collect multiple blood pressure measurements and information on hypertension diagnosis and treatment at each survey wave. The surveys employed different sampling procedures to recruit participants. The IFLS, MxFLS and NIDS recruited a random sample of households, where all (MxFLS, NIDS) or a predefined number (IFLS) of household members were interviewed. The CHNS randomly selected four counties from each of the nine Chinese provinces and then selected households (all members were interviewed) from a random sample of primary sampling units within counties. Appendix I provides more detailed information on the sampling designs for each survey.

For each country, we follow cohort members across two survey waves approximately seven years apart (CHNS 2009-2015, IFLS 2007-2014, MxFLS 2005/6-2009/12, NIDS 2008-2017), which we label chronologically as the baseline and follow-up waves. We chose the most recently available follow-up wave and the baseline wave for each survey to maximise temporal comparability between countries. We restrict our study to adults aged 40 years and older in the baseline wave, since the WHO PEN guidelines recommend hypertension screening and treatment from this age.^12^ We further limit our sample to individuals with non-missing information on blood pressure measurements and hypertension diagnosis and treatment history at both baseline and follow-up. Information on sex, schooling, and whether household was located in an urban or rural area in the baseline wave was also necessary for inclusion in the analysis. All variables of interest and anthropometric measurements were collected by trained field staff in structured interviews in a face-to-face setting and answered either exclusively by the head of household and their spouse, or by all members of a household.

### Measurements

#### Defining hypertension

Blood pressure was measured in each survey by trained field staff with readings taken on seated individuals in specified time intervals with either a manual mercury sphygmomanometer (CHNS) or digital self-inflating sphygmomanometers (IFLS, MxFLs, NIDS). Individuals in the MxFLS and NIDS were measured twice in each wave while those in the CHNS and IFLS three times. To reduce measurement error and mitigate bias from the white-coat effect^13^, we took the average of the second two BP measurements in the CHNS and IFLS and the average of the two measurements in the MxFLS and NIDS. Further details on the measurement conditions in each survey can be found in Appendix I. In addition to blood pressure measurements, individuals were asked if they had ever been diagnosed with elevated blood pressure by a health care professional and whether they were currently taking anti-hypertensive medication (among those that said yes to having been diagnosed). Following the 2020 International Society for Hypertension Guidelines^14^, we classified individuals as hypertensive if they met any of the following conditions: mean systolic blood pressure (SBP) ≥ 140 mmHg, mean diastolic blood pressure (DBP) ≥ 90 mmHg, or a reported prior diagnosis of hypertension.

#### Stages of the hypertension care continuum

Based on an individual’s measured BP, diagnosis, and treatment status, we classified individuals into the following four mutually exclusive continuum of care stages at both baseline and at follow-up (Figure in Appendix VI): undiagnosed; diagnosed and untreated; diagnosed, treated, but uncontrolled (SBP ≥ 140 mmHg or DBP ≥ 90 mmHg); diagnosed, treated, and controlled (SBP < 140 mmHg and DBP < 90 mmHg). We restricted our sample to those who were hypertensive at baseline. A small group of individuals were undiagnosed at baseline with a SBP ≥ 140 mmHg or DBP ≥ 90 mmHg and were still undiagnosed but had a measured SBP < 140 mmHg and DBP < 90 mmHg at follow-up. We classified these individuals as indeterminate at follow-up.

#### Social and demographic variables

We used information on each respondent’s baseline self-reported age, sex, household location (the definitions used by each of the surveys to delimit rural/urban household location are reported in Appendix II), and highest level of attained education. Since the countries have different national education systems, we classified schooling into the following comparable categories using the International Standard Classification of Education^15^: unschooled, primary, secondary, and tertiary education (Appendix III shows the specific schooling levels used in each category by country).

### Statistical analyses

This is a complete case analysis. We first describe individuals’ transition through the continuum over time. Starting from each baseline stage, we estimate the probabilities of progressing forward to any subsequent continuum stage or regressing backward to a prior continuum stage between the baseline and follow-up waves. Next, we estimate the probability of four important clinical transitions (all between the baseline and endline waves): becoming diagnosed (among all those who are undiagnosed at baseline); becoming treated (among all those who are untreated at baseline); stopping treatment (among all those who report taking treatment at baseline); and reaching the diagnosed, treated, and controlled stage (among all those at prior stages at baseline).

Lastly, we investigate if certain social and demographic subgroups are at an increased risk of not moving forward through the continuum or moving backwards to prior stages. We consider differences based on age group (40 – 60 years old or 60 +), sex (female or male), schooling (unschooled, primary, secondary, or tertiary), and household location (urban or rural). To estimate the magnitude of differences, we use Poisson regression models for progression and regression through the cascade with each sociodemographic characteristic as the main exposure (separately for each exposure). Importantly, to adjust for the differential baseline distribution of individuals across the continuum stages, we also include indicator variables for each baseline stage in all regressions. We estimate Poisson rather than logistic regression models so that our main coefficient estimates are expressed as risk, rather than odds, ratios.

All analyses are weighted to be nationally representative, except those performed for China due to the unavailability of weights for the CHNS survey.

### Ethical approval

This study was exempt from institutional review board approval because the data are publicly available and deidentified.

## RESULTS

### Sample sizes and missingness

We extracted all individuals aged 40 and over from the baseline samples for each survey (China: 7380, Indonesia: 14304, Mexico: 10096, South Africa: 7606). Among our age-eligible samples, 1% of individuals were missing sociodemographic information, while 16-26% (China: 16%, Indonesia: 25%, Mexico: 24%, South Africa: 26%) were missing information on BP measurements, treatment, or diagnosis information at baseline (Figure 1). After excluding individuals with missing information at baseline, we excluded 28-52% (China: 52%, Indonesia: 39%, Mexico: 49%, South Africa: 28%) of individuals because they were not hypertensive at baseline and thus did not meet the study inclusion criteria. At follow-up, sources of missing information included loss to follow-up (China: 2%, Indonesia: 8%, Mexico: 6%, South Africa: 12%), death between waves (China: 6%, Indonesia: 16%, Mexico: 7%, South Africa: 25%), and incomplete collection of anthropometric parameters resulting in missing blood pressure measurements and/or related medical history at follow-up (China: 34%, Indonesia: 8%, Mexico: 14%, South Africa: 16%).

**Figure 1.**
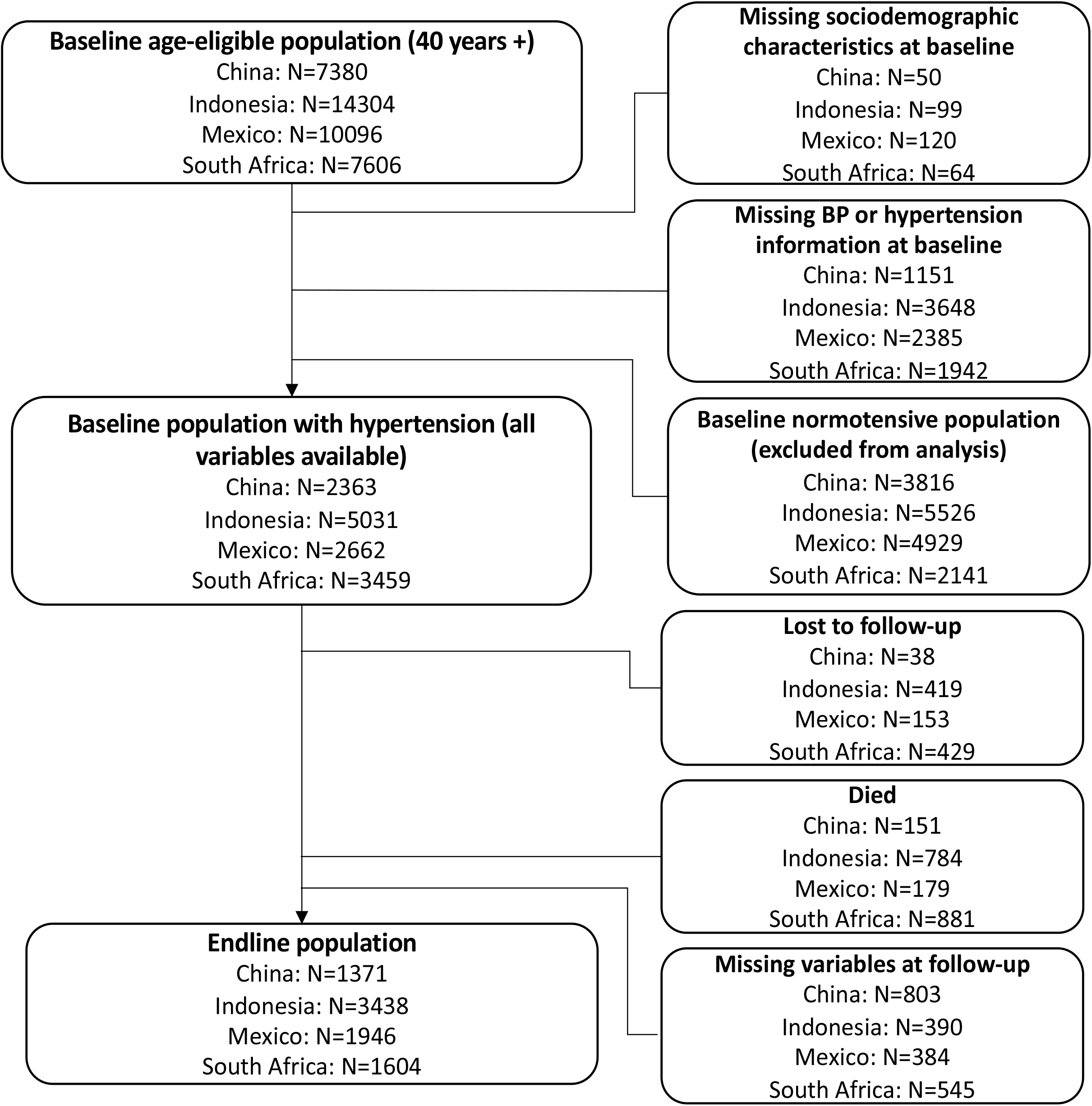
Flow diagram of sample selection from baseline to endline

Compared to our age-eligible population, the baseline population of individuals with hypertension (after excluding those who were missing baseline information) was slightly older (China: Mean: 61·0, SD: [11·3] vs. 56·9 [11·4], Indonesia: 56·5 [11·4] vs. 55·1 [11·8], Mexico: 58·3 [11·9] vs. 55·9 [12·3], South Africa: 57·7 [11·7] vs. 55·7 [11·9]) which is to be expected considering the increase in prevalence of hypertension with age.^16^ The two populations were similar in terms of sex distribution, household location and educational attainment with slightly higher proportions of females (Indonesia: 58% vs. 53%, Mexico: 59% vs. 53%, South Africa: 67% vs, 60%) and primary (Mexico: 57% vs. 52%), or no education (China: 35% vs. 29%) in the hypertensive baseline samples. There were no stark age and sex differences between our baseline hypertensive population and individuals excluded at follow-up due to missing information, although the latter were more likely to be from an urban household across all countries (China: 45% vs. 35%, Indonesia: 63% vs. 53%, Mexico: 66% vs. 57%, South Africa: 59% vs. 49%). There were also no apparent sociodemographic dissimilarities between our baseline hypertensive population and the final analytical samples, aside a slightly higher proportion of females (South Africa: 73% vs. 67%) and a lower proportion of urban households in our final analytic samples (China: 30% vs. 35%, South Africa: 44% vs. 49%) in the countries listed. A detailed summary of the sex, age, and urban household composition of the age-eligible samples compared to our baseline hypertensive populations and to the final analytic samples can be found in Appendix IV.

### Sample characteristics at baseline

Table 1 provides weighted descriptive characteristics for each of the country samples at baseline (unweighted for China due to the lack of survey weights). Mean age was highest in China (59·1, SE: 0·27), but above 50 years across all countries (Indonesia 53·9, SE: 0·18, Mexico: 57·0, SE: 0·38, South Africa: 54·4, SE: 0·39). The proportion of females in our samples were 51·7% in China, 57·9% in Indonesia, 61·9% in Mexico, and 67·9% in South Africa. Approximately half of the sample population live in urban households in all countries but China, which has predominantly rural households (69·9%). The proportion of individuals with no education varied from 17·5% in Indonesia to 32% in China, while the proportion with a tertiary education was below 10% in all countries.

**Table 1.**
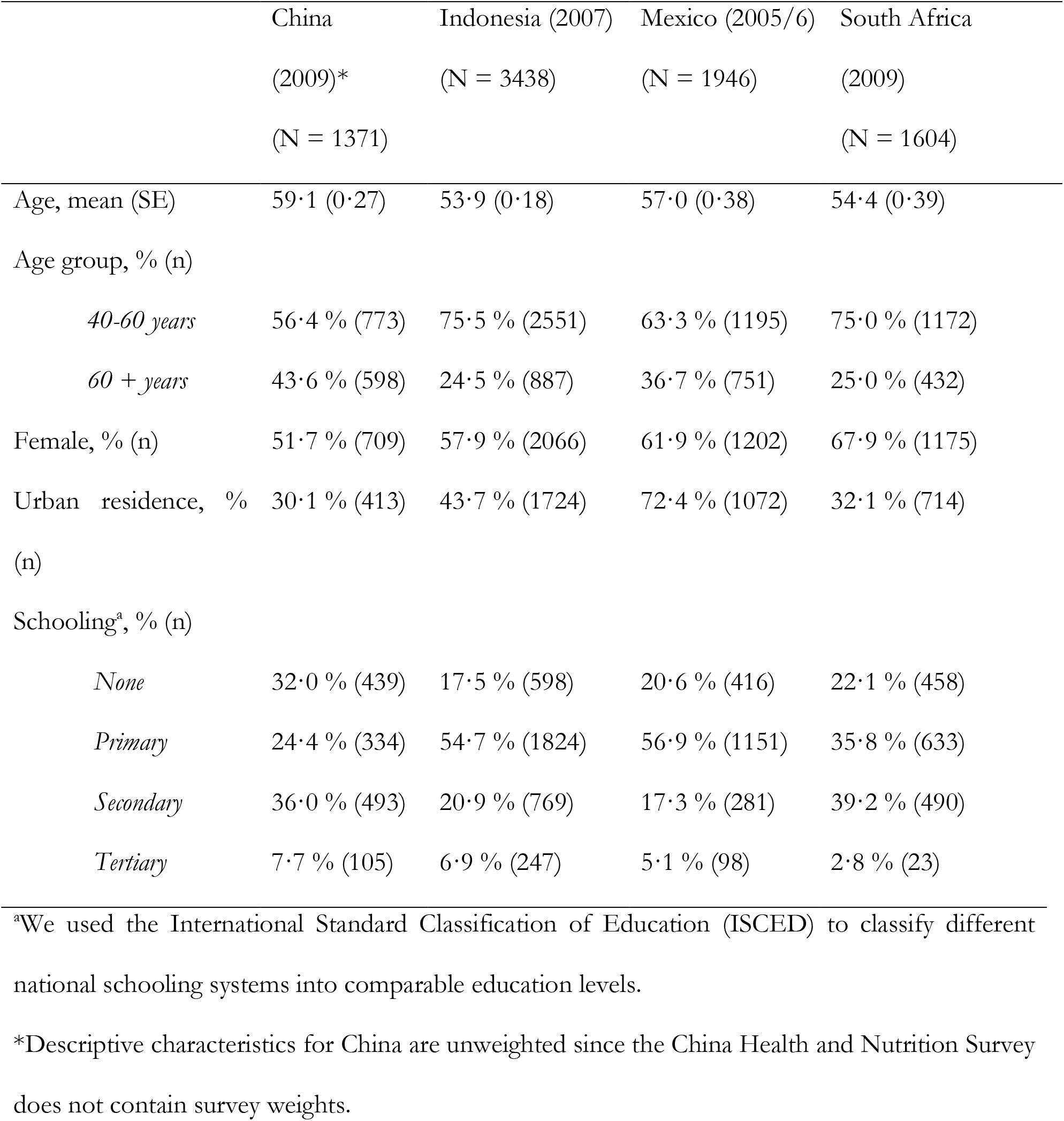
Baseline weighted descriptive characteristics for each country sample

|  | China<br>(2009)*<br>(N = 1371) | Indonesia (2007)<br>(N = 3438) | Mexico (2005/6)<br>(N = 1946) | South Africa<br>(2009)<br>(N = 1604) |
| --- | --- | --- | --- | --- |
| Age, mean (SE) | 59.1 (0.27) | 53.9 (0.18) | 57.0 (0.38) | 54.4 (0.39) |
| Age group, % (n) |  |  |  |  |
| *40-60 years* | 56.4 % (773) | 75.5 % (2551) | 63.3 % (1195) | 75.0 % (1172) |
| *60 + years* | 43.6 % (598) | 24.5 % (887) | 36.7 % (751) | 25.0 % (432) |
| Female, % (n) | 51.7 % (709) | 57.9 % (2066) | 61.9 % (1202) | 67.9 % (1175) |
| Urban residence, %<br>(n) | 30.1 % (413) | 43.7 % (1724) | 72.4 % (1072) | 32.1 % (714) |
| Schooling<sup>a</sup>, % (n) |  |  |  |  |
| *None* | 32.0 % (439) | 17.5 % (598) | 20.6 % (416) | 22.1 % (458) |
| *Primary* | 24.4 % (334) | 54.7 % (1824) | 56.9 % (1151) | 35.8 % (633) |
| *Secondary* | 36.0 % (493) | 20.9 % (769) | 17.3 % (281) | 39.2 % (490) |
| *Tertiary* | 7.7 % (105) | 6.9 % (247) | 5.1 % (98) | 2.8 % (23) |

<sup>a</sup>We used the International Standard Classification of Education (ISCED) to classify different national schooling systems into comparable education levels.
\*Descriptive characteristics for China are unweighted since the China Health and Nutrition Survey does not contain survey weights.

### Hypertension care continuum at baseline

The majority of individuals with hypertension at baseline were undiagnosed (Figure 2, China: 57% [95% CI 54-59%], Indonesia: 65% [95% CI 63-66%], Mexico: 52% [95% CI 49-56%], South Africa: 53% [95% CI 49-57%]). Fewer than one in six individuals were diagnosed and untreated (China: 9% [95% CI 7-10%], Indonesia: 11% [95% CI 10-12%], Mexico: 15% [95% CI 12-18%], South Africa: 8% [95% CI 6-10%]), and approximately one in four were diagnosed and treated with uncontrolled BP (China: 25% [95% CI 23-27%], Indonesia: 17% [95% CI 16-18%], Mexico: 15% [95% CI 13-17%], South Africa: 22% [95% CI 19-25%]). Among all individuals with hypertension, less than 20% had a controlled BP at baseline, with especially low proportions in China and Indonesia (Mexico: 18% [95% CI 15-20%], South Africa: 17% [95% CI 13-20%], China: 9% [95% CI 8-11%], and Indonesia: 7% [95% CI 6-8%]).

**Figure 2.**
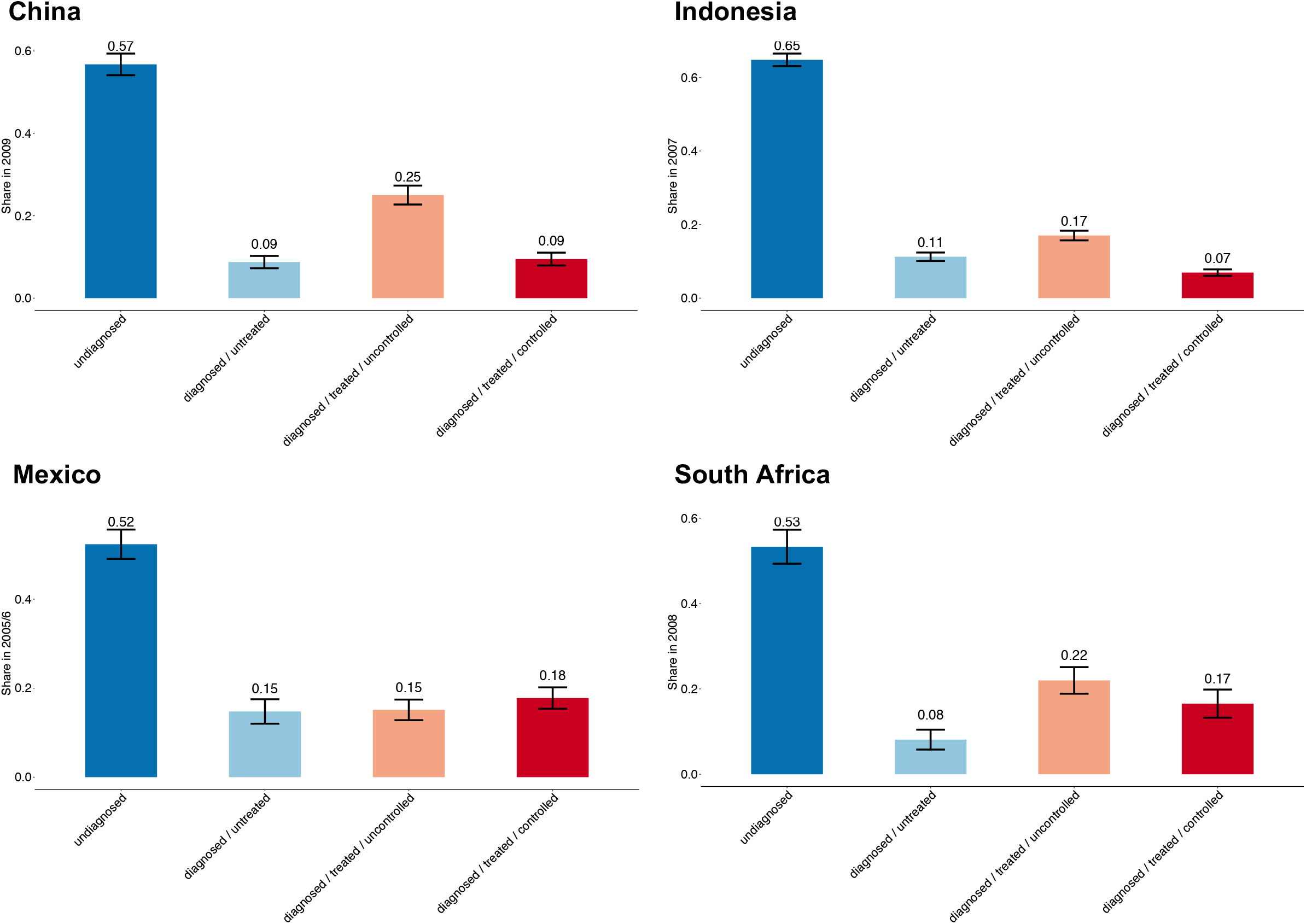
Distribution in the continuum of care for hypertension at baseline ^a^Survey years: China (2009 to 2015), Indonesia (2007 to 2014), Mexico (2005/6 to 2009/12), South Africa (2008 to 2017). ^b^We weighted estimates in Indonesia, Mexico, and South Africa to be nationally representative; the China Health and Nutrition Survey does not contain survey weights.

### Probability of progression and regression by baseline continuum stage

From each baseline stage and across all countries, there was a less than 50% probability that an individual progressed to a more advanced continuum stage (Figure 3). For example, for those that are undiagnosed at baseline, the probability of progressing to a later stage of the continuum was 30% in China (95% CI 26-33%), 30% in Indonesia (95% CI 28-32%), 27% in Mexico (95% CI 24-30%), and 37% in South Africa (95% CI 34-40%). Importantly, the probability of regressing to a prior stage was also high across all countries and continuum stages. For example, for those who were diagnosed, treated, but uncontrolled at baseline, the probability of regressing backwards ranged from 65% (95% CI 62-69%) in Indonesia to 35% (95% CI 30-40%) in China, 27% (95% CI 22-32%) in Mexico (27%), and 25% (95% CI 21-29%) in South Africa. The most striking finding, however, is the high probability of regression from a baseline status of blood pressure control to prior stages of the continuum across all countries, with the highest regression probability observed in Indonesia (92% [95% CI 88-95%]), followed by Mexico (77% [95% CI 72-81%]), China (76% [95% CI 68-83%]), and South Africa (48% [95% CI 42-54%]).

**Figure 3.**
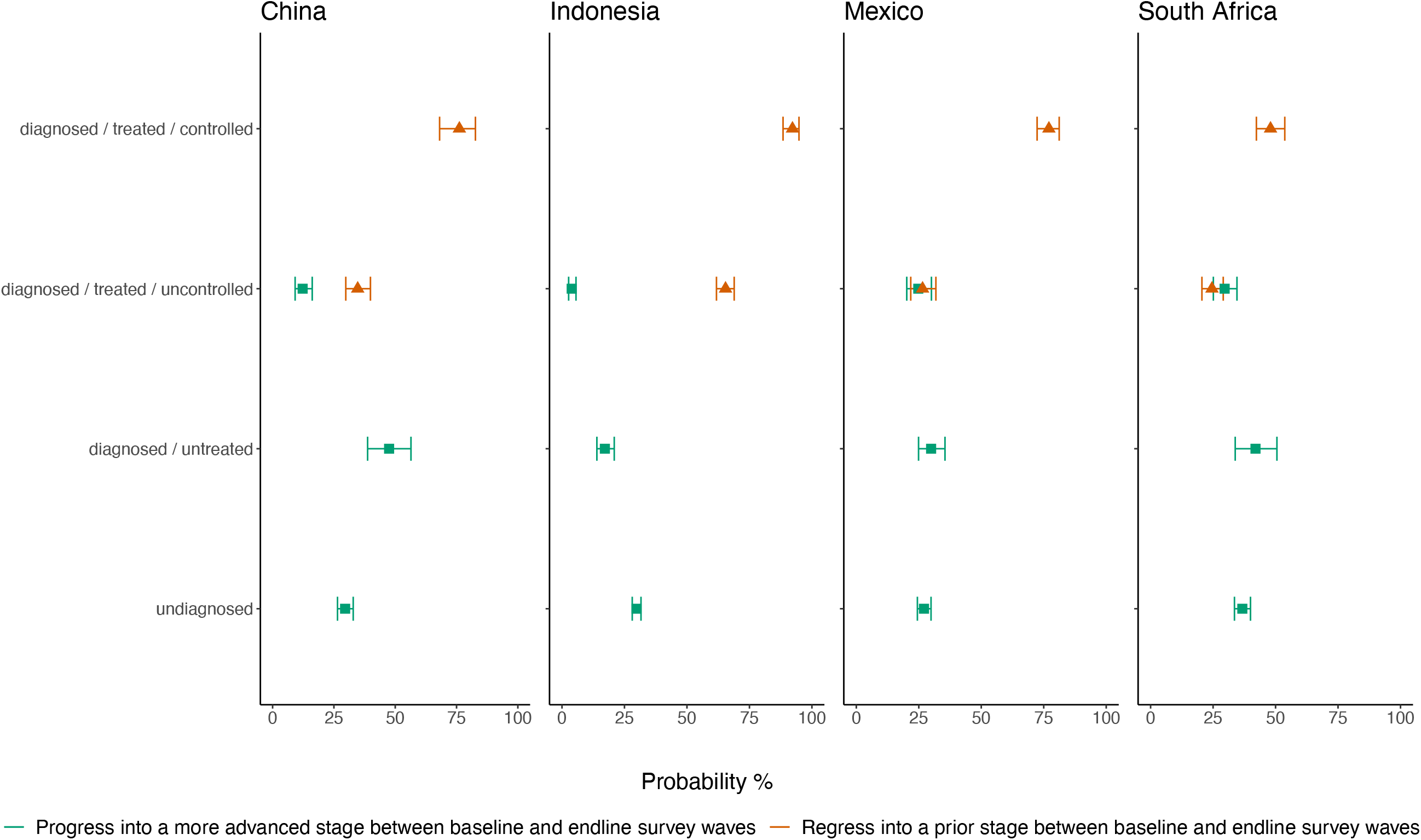
Probability of care progression and regression between baseline and endline survey waves based on baseline continuum stage ^a^We defined progression as the movement of individuals from their baseline stage into a more advanced stage of the hypertension care continuum between survey waves. Once blood pressure control is achieved, no further progression along the continuum is possible. ^b^We defined regression as the backward movement of individuals from their baseline stage into a prior stage of the hypertension care continuum between survey waves. Once study subjects have been diagnosed, they can no longer be undiagnosed. ^c^ Survey years: China (2009 to 2015), Indonesia (2007 to 2014), Mexico (2005/6 to 2009/12), South Africa (2008 to 2017).

### Probability of important clinical transitions

Just over one in four individuals who were undiagnosed at baseline were diagnosed at the follow-up survey wave (Figure 4, China 30% [95% CI 26-33%], Indonesia 30% [95% CI 28-32%], Mexico 27% [95% CI 25-29%], South Africa 37% [95% CI 34-39%]). The probability of becoming treated among those who were untreated at baseline was 17% [95% CI 14-21%] in Indonesia, 30% [95% CI 28-32%] in Mexico, 48% [95% CI 39-56%] in China, and 42% [95% CI 40-44%] in South Africa. Among those with uncontrolled blood pressure at baseline, the probability of reaching treated and controlled blood pressure at endline was 21% (95% CI 19-23%) in South Africa, 9% (95% CI 7-11%) in China, and 11% (95% CI 10-13%) in Mexico, and just 2% (95% CI 1-2%) in Indonesia. Importantly, there was also a high probability of stopping treatment between the baseline and endline waves across all four countries (China 36% [95% CI 32-41%], Indonesia 70% [95% CI 67-73%], Mexico 34% [5% CI 32-36%], South Africa 25% [23-27%]).

**Figure 4.**
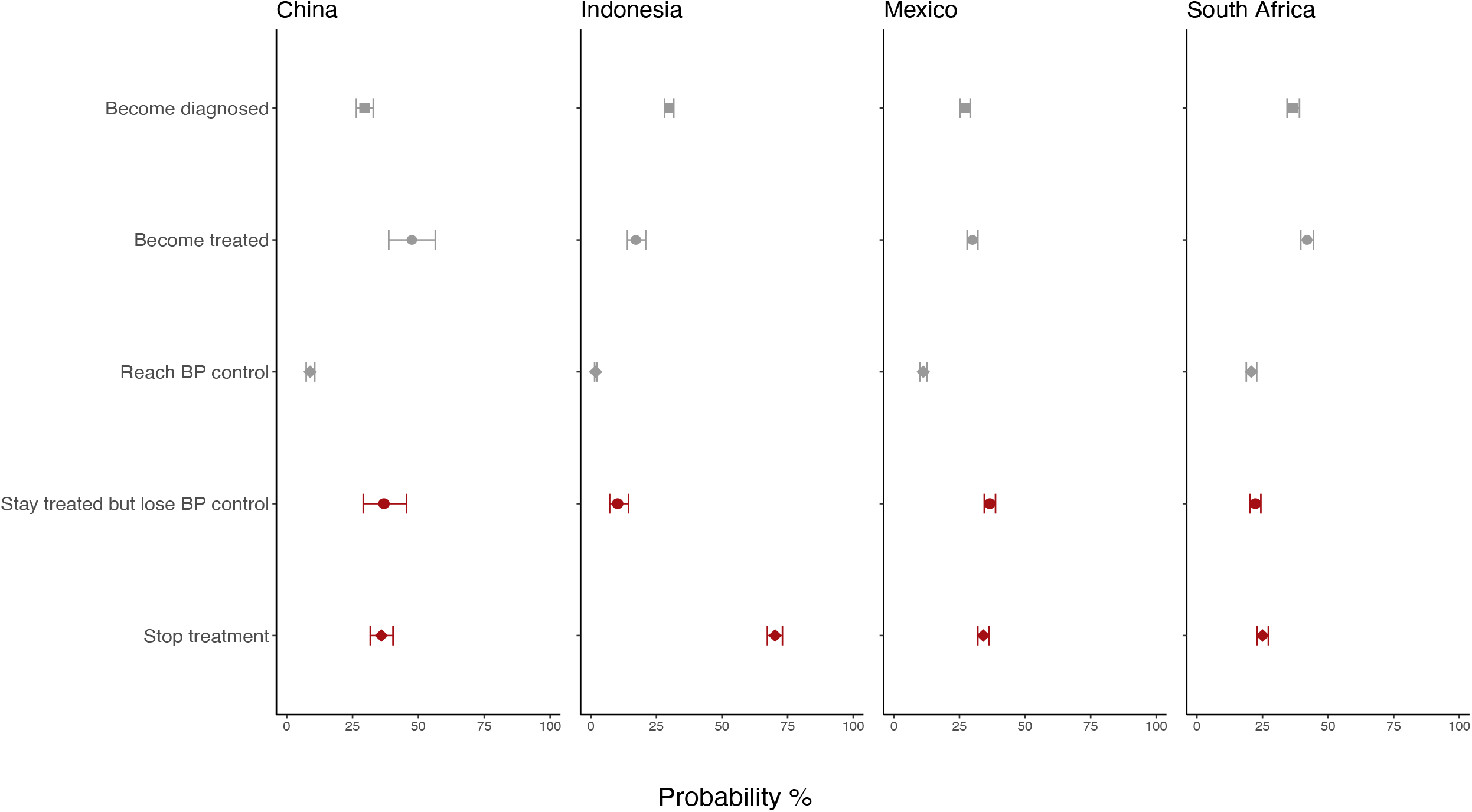
Probability of diagnosis, treatment, blood pressure control and treatment discontinuation based on baseline continuum stage ^a^We defined “Become diagnosed” as the movement of individuals from being undiagnosed at baseline to any stage of diagnosed hypertension inside the hypertension care continuum. ^b^We defined “Become treated” as the movement of individuals from any baseline stage of untreated hypertension into any stage of treated hypertension. ^c^We defined “Reach BP control” as the movement of individuals from any baseline stage of uncontrolled hypertension to a stage of treated, controlled BP. ^d^We defined “Stay treated but lose BP control” as the movement of individuals from a baseline stage of treated, controlled hypertension to a stage of treated, uncontrolled hypertension. ^e^We defined “Stop treatment” as the backward movement of individuals from baseline treated hypertension into any prior stage of untreated hypertension. ^f^Survey years: China (2009 to 2015), Indonesia (2007 to 2014), Mexico (2005/6 to 2009/12), South Africa (2008 to 2017)

### Movement in the care continuum stratified by sociodemographic characteristics

Females were more likely to progress to a more advanced continuum stage than males in Indonesia and Mexico (Indonesia: RR 1·50 [95% CI 1·30-1·72], p<0·001, Table 2); Mexico: RR 1·30 [95% CI 1·10-1·54], p = 0·003), with weaker evidence of a female advantage in South Africa (RR: 1·12 [0·96-1·32], p = 0·148), and no difference between sexes in China (RR 0·99 [95% CI 0·79-1·22], p=0·897). Compared to individuals aged 60+, younger adults were less likely to progress in China (RR: 0·74 [95% CI 0·59-0·92], p=0·006) and South Africa (RR: 0·81 [95% CI 0·69-0·95], p=0·009). We found evidence of a rural disadvantage in all countries but China, where rural individuals were less likely to progress in Indonesia (RR: 0·82 [95% CI 0·72-0·94], p=0·003) and Mexico (RR: 0·79 [95% CI 0·66-0·96], p=0·020), and more likely to regress backwards in South Africa (RR: 1·36 [95% CI 1·10-1·67], p = 0·004). In three out of four countries, individuals with a higher level of education were more likely to progress or less likely to regress than people with lower education. This trend is evident in Mexico, where having a secondary (RR 0·74 [95% CI 0·57-0·94], p=0·015) or tertiary (RR 0·71 [95% CI 0·47-1·02], p=0·079) education was associated with a significantly lower risk of regressing compared to unschooled individuals. Similarly, in South Africa, having a tertiary degree conferred a regression advantage (RR 0·15 [95% CI 0·03-0·43], p=0·002). In Indonesia, both secondary (RR 1·24 [95% CI 1·01-1·53], p=0·037) and tertiary education (RR 1·30 [95% CI 0·99-1·70], p=0·053) were positively associated with progressing through the continuum.

**Table 2.**
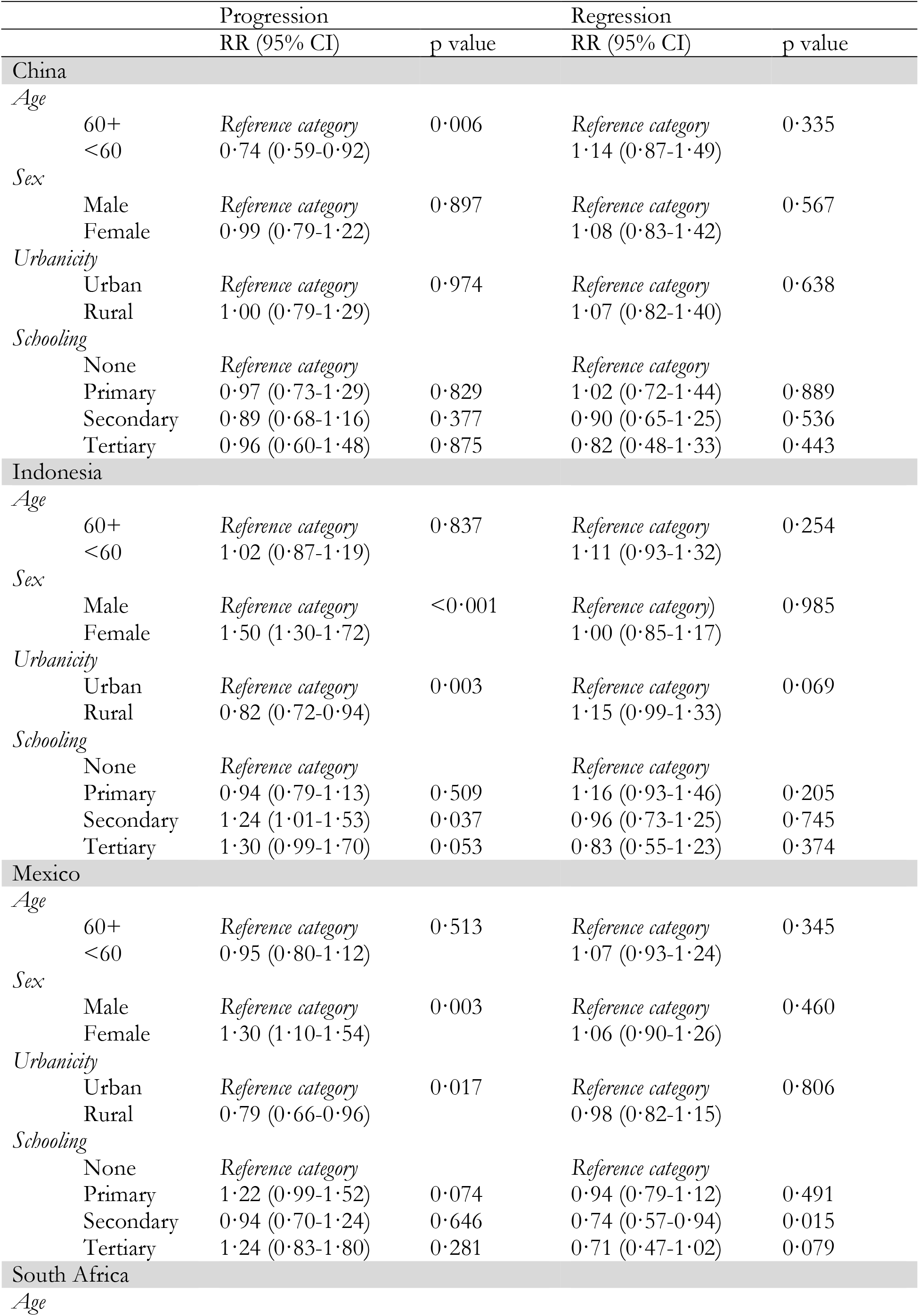
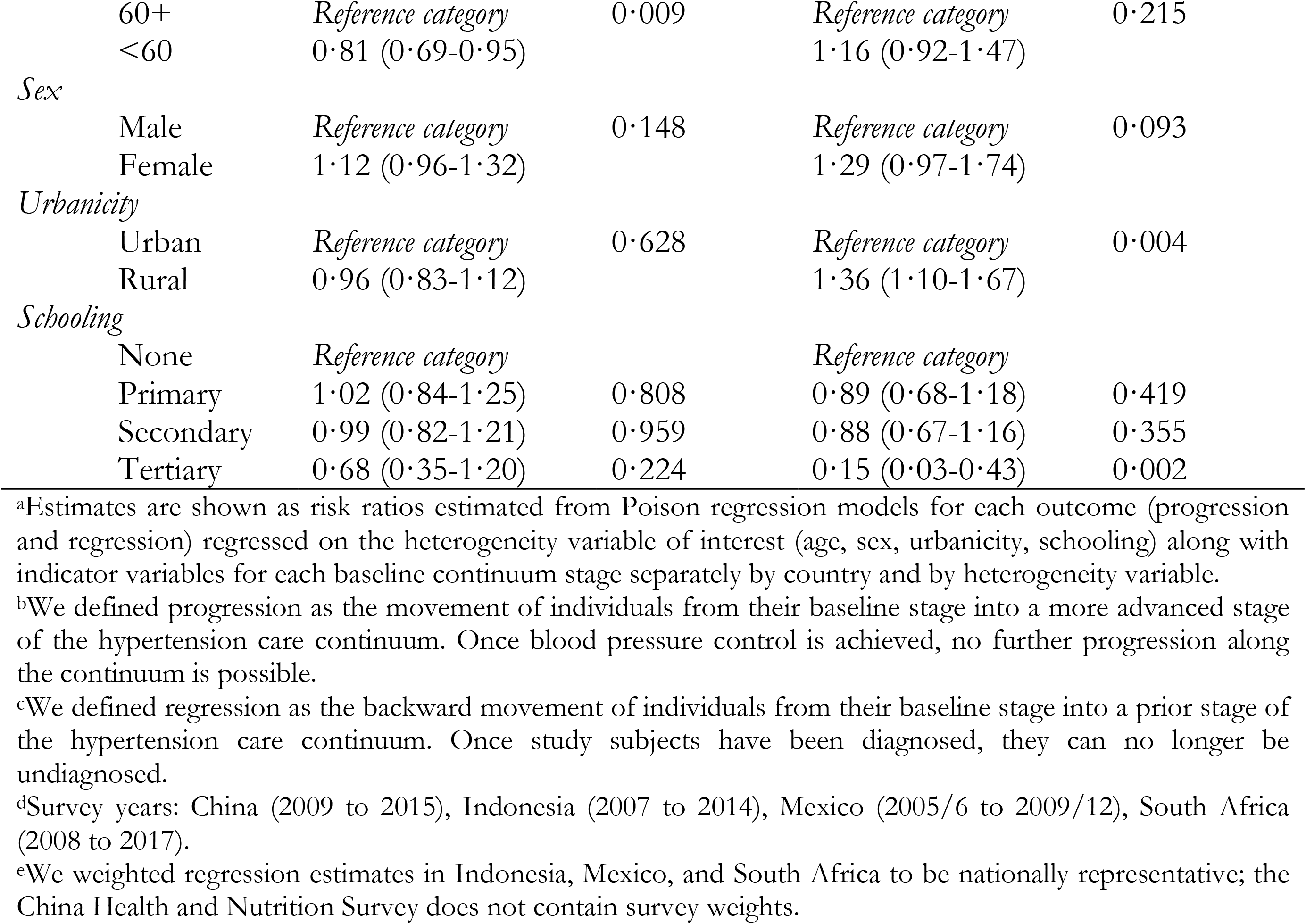
Age, sex, urbanicity, and schooling differences in the relative risk of progressing and regressing on the hypertension care continuum

## DISCUSSION

In this longitudinal analysis of the hypertension care continuum in four of the world’s most populous middle-income countries, we found that over a five to nine-year period, up to nine in ten individuals with treated and controlled blood pressure do not maintain control and up to two-thirds of individuals taking treatment discontinue it over time. There was also significant stasis at the early stages of the care continuum: three of four undiagnosed individuals remain undiagnosed five to nine years later and among those individuals who are diagnosed but not taking treatment, nearly all remain untreated at endline. This longitudinal perspective reveals that achievement of BP control is seldom sustained over time and that policies solely aimed at improving diagnosis or initiating treatment may not lead to large improvements in control, as those who are diagnosed are unlikely to start treatment and those who do start treatment tend to discontinue treatment over time.

Existing studies largely associate poor treatment rates to barriers in access to diagnostic services and a failure to initiate treatment among those who are diagnosed.^17,18,19,20^ However, we find that aside low initiation, treatment discontinuation is an equally important contributor to the large observed hypertension treatment gaps. This dual contribution to suboptimal treatment is important for policy efforts since the reasons for treatment discontinuation likely diverge from those explaining non-initiation of treatment. Inadequate treatment initiation has been linked to limitations in access to primary and community health care services, affordability, inequitable distributions of facilities, and trained health workers.^21,22,23,24,25^ Patients’ socioeconomic background and level of health literacy^26^, multi-drug treatment plans, and the need for multiple titration visits have been implicated in treatment non-adherence.^27^ In contrast, there are few large-scale studies exploring the causes of widespread treatment discontinuation in LMICs. Based on a sample of adults with hypertension in Chennai, India, Sudharsanan et al. find that treatment discontinuation may be linked to the false belief that blood pressure medications can be discontinued once BP control has been achieved.^28^ Davies et al. describe similar attitudes in a population of adults with hypertension in Sierra Leone, drawing a parallel across two continents.^29^

Cross-sectional continuums convey the impression that those in the final treated and controlled stage have achieved ideal hypertension care and that health policy should focus on bringing individuals at other stages to this point. Yet, we observe that this last stage is not permanent, and the majority of people lose BP control and discontinue treatment over time. This result is consistent with a health record-based study in the UK of over 150 000 individuals with newly diagnosed hypertension, which shows that only 0·6% of people in the study sample are able to sustain a year-round controlled BP in the first year of follow-up.^30^ Loss of BP control may reflect inadequate follow-up practices and a failure by service providers to intensify treatment over time in patients with physiologically progressive hypertension. As such, there is a crucial need to move beyond cross-sectional monitoring of hypertension care performance and to untangle the specific reasons behind treatment discontinuation and loss of BP control^31^ in these settings.

South Africa emerged as the country with the lowest rates of treatment discontinuation and blood pressure control loss, as well as the highest probabilities of individuals reaching blood pressure control overall and across different sociodemographic subgroups. These findings diverge from those in other countries in our analysis and may be due to the country-specific context. In 2013, South Africa started implementing the ‘Ideal Clinic’ programme to systematically improve the quality of care delivered in Primary Health Care facilities in view of achieving Universal Health Coverage through a novel National Insurance model.^32^ It is possible that the participants in our surveys benefitted from better chronic disease care during the follow-up period between waves. South Africa also has the largest HIV-positive population in the world, with over seven million individuals and over 20% of the adult population affected.^33^Substantial progress has been made in reducing HIV-related mortality and increasing antiretroviral (ART) coverage.^34,35^ Recent studies show that use of antiretroviral agents is significantly associated with greater awareness and treatment of hypertension in the HIV-positive population.^36^ Patients treated with ART also appear to have considerably better blood pressure control if their viral loads are undetectable.^37^ This implies that established ART programmes may serve as platforms for the delivery of cardiometabolic care and the generalised adoption of chronic disease management behaviours, which facilitate the attainment of long-lasting treatment compliance and disease control.^38,39^ Alternatively, in our study, South Africa also had the largest number of individuals who died between survey waves; therefore, these findings could also reflect selective mortality among those less likely to progress through the continuum.

In all countries but China, women were more likely to progress through the care continuum. This finding is consistent with previous literature reporting a better control of hypertension in women^8,40^ and a tendency to seek out medical care more frequently than men when confronted with the same disease.^41^ At younger ages, women are more regularly brought into contact with health systems than men for reproductive and birth-related care.^42^ This may favour the normalisation of health-seeking behaviour in women which persists over time into their post-reproductive ages. We also observed individuals from rural households had a disadvantage in progressing and regressing through the hypertension care continuum compared to urban ones in three out of four countries. Similar effects have been described in large cross-sectional continuums like the Prospective Urban Rural Epidemiology (PURE) study, in which rates of awareness, treatment, and control of hypertension were significantly lower in rural compared to urban communities across seven LMICs.^9^ Finally, people with a secondary or tertiary level education tended to either regress less or be more likely to progress within the continuum. This finding is consistent with results from another cross-sectional study of 1·1 million adults from 44 LMICs, which showed secondary or higher educational attainment was positively associated with reaching every step of the continuum;^8^ however, due to the small proportion of people with tertiary education in our country samples, conclusions concerning this group should be drawn with caution.

Our study has a number of limitations. First, a substantial share of individuals was excluded due to missing data or loss to follow-up. These individuals were more likely to be over 60 years old, male, and from an urban household. Given this, our findings may not represent the care patterns among the overall population and should be interpreted as being most relevant to populations similar in composition to our study sample. Second, the classification of continuum stages is based on self-reported diagnosis and treatment information which may not always be accurate. Indeed, we observed approximately 30% of baseline diagnosed individuals reporting never having been diagnosed at endline. We also did not apply lower blood pressure thresholds when classifying diabetic individuals as hypertensive as biomarker-based information on diabetes status was not available in the NIDS. Third, a minority of individuals with undiagnosed hypertension present with blood pressure in normal ranges at follow-up. While an improvement in blood pressure may be the result of self-directed lifestyle changes, the discrepancies between baseline and follow-up may be due to limitations in the diagnostic accuracy of ambulatory measurements. In clinical practice, a hypertension diagnosis is based on multiple elevated blood pressure measurements, since ambulatory settings are susceptible to physiological fluctuations and white-coat hypertension.^43^ We attempted to limit these effects by using the mean of two consecutive blood pressure measurements, which were taken in pre-defined time intervals and in a seated position. Nevertheless, the potential measurement bias may have led us to misclassify the population of individuals with undiagnosed hypertension. Lastly, we focus on changes in hypertension care status between two time points separated by seven years on average and do not have information on how many people initiated and discontinued treatment during this interval. It is also important to note that individuals with baseline undiagnosed hypertension become aware of having a high BP at the time of interview, which may render them more likely to seek out a diagnosis than someone in the general population during the follow-up period. Both these limitations, however, imply that our current estimates of diagnostic gaps, treatment discontinuation, and loss of blood pressure control are conservative.

Despite these limitations, to our knowledge, this is the first study of longitudinal hypertension care continuums with nationally representative data from an LMIC. Our analysis includes four large middle-income countries which cover close to one-fourth of the world’s population. We find that diagnosis and treatment initiation are major bottlenecks, which must be tackled concurrently to avoid individuals dwelling in one stage and to facilitate movement along the care continuum. Nonetheless, treatment discontinuation and loss of BP control are equally problematic and reveal the dynamic nature and long-lasting demands of chronic disease care.

## Data Availability

The data employed are publicly available and freely downloadable at the respective websites of the China Health and Nutrition Survey (CHNS), the Indonesian Family Life Survey (IFLS), the Mexican Family Life Survey (MxFLS) and the South African National Income Dynamics Study (NIDS). All code used for data analysis will be made available on the corresponding author's website.

## Authors’ contributions

NS and NM conceived the study. NS and NM had full access to the data, led the data analysis, and take responsibility for the integrity and accuracy of the analysis. NS and NM wrote the first draft of the manuscript, and all authors provided crucial input to the drafting of the final manuscript. All authors have approved the final version.

## Acknowledgements

Financial support was provided to NS, NM and PS by the Alexander von Humboldt Foundation, by the Else Kröner-Fresenius Foundation and by the European Research Council, respectively.

