## Supplemental Material for "Progress, stasis, and regression through the hypertension care continuum: Longitudinal evidence from population-based cohort data in four populous middle-income countries"

### SUPPLEMENTARY MATERIAL: APPENDICES

**Appendix I:** Survey sample designs
**Appendix II:** Definitions of hypertension diagnosis/treatment and urbanicity for each survey
**Appendix III:** Classification of national education systems
**Appendix IV:** Comparison of sample composition after exclusion of missing data
**Appendix V:** Transition probabilities between continuum stages for each country
**Appendix VI:** Successive stages of the hypertension care continuum

### Appendix I: Sampling designs used in each survey

| Survey name | Years of data collection used in study | Eligibility criteria | Sample design | Blood pressure measurements/ Eligibility |
|---|---|---|---|---|
| China Health and Nutrition Survey | 2009, 2015 | All households | *First stage*: random sample of 4 counties from each of 9 provinces<br>*Second stage*: random sample of 360 PSUs<br>*Third stage*: random sample of HHs<br>*Interviews:* all HH members | 3 BP measurements, mercury sphygmomanometer/ Children age 7 and older and all adults |
| Indonesian Family Life Survey | 2007, 2014/15 | All households | *First stage*: random sample of 321 census enumeration areas from 13 provinces<br>*Second stage*: random sample of 20-30 HHs<br>*Interviews*: HH head and spouse, two randomly selected children, a randomly selected individual age 50+ and their spouse, in 25% of randomly selected HHs an individual aged 15-49 and their spouse | 3 BP measurements, Omron digital self-inflating sphygmomanometer/ Children age 15 and older and all adults |
| Mexican Family Life Survey | 2005/6, 2009/12 | All households | *First stage:* nationally/regionally representative PSUs selected based on pre-established demographic/economic variables<br>*Second stage*: random sample of HHs<br>*Interviews*: all HH members | 2 BP measurements/ Children age 15 and older and all adults |
| South African National Income Dynamics Study | 2008, 2017 | All households | *First stage:* random sample of 400 PSUs from Stats SA Master Sample<br>*Second stage*: random sample of dwellings and identification of eligible HHs<br>*Interviews*: all HH members | 2 BP measurements/ Children age 15 and older and all adults |

**Appendix II: Definitions of hypertension diagnosis/treatment and urbanicity for each survey**

**China Health and Nutrition Survey (2009, 2015)**

- Definition of hypertension diagnosis/treatment:

- Has a doctor ever told you that you suffer from high blood pressure? (Yes/No/unknown)
- Are you currently taking anti-hypertension drugs? (Yes/No/unknown)

- Definition of urban vs. rural household:

- Urban = city, town or county capital city
- Rural = suburban or rural village

**Indonesian Family Life Survey (2007, 2014)**

- Definition of hypertension diagnosis/treatment:

- Have a doctor/paramedic/nurse/ midwife ever told you that you had hypertension? (Yes/No)
- In order to deal with [...] are you currently taking prescribed medication on a weekly basis? (Yes/No)

- Definition of urban vs. rural household:

The urban/rural code is based on the BPS code, which is assigned to each locality by the Indonesian Bureau of Statistics (BPS) according to indicators (population density, share of population in agriculture, percentage of area occupied by non-housing buildings, infrastructure indicators (access to electricity/telephone, hospitals, banks, schools, roads, markets, hotels, cinemas etc.)). The BPS can be contacted for a detailed, up-to-date list of indicators and scoring system used.

**Mexican Family Life Survey (2005/6, 2009/12)**

- Definition of hypertension diagnosis/treatment:

- Have you ever been diagnosed with hypertension? (Yes/No)
- Currently, do you take medicine for this illness on a regular basis? (Yes/No)

- Definition of urban vs. rural household:

- Urban areas:

Estrato=1 Households located in localities with more than 100 000 inhabitants.
Estrato=2 Households located in localities with populations between 15 000 and 100 000.
Estrato=3 Households located in towns with a population between 2 500 and 15 000.

- Rural Areas:

Estrato=4 Households located in areas with less than 2 500 inhabitants.

**South Africa National Dynamics Income Study (2008, 2017)**

- Definition of hypertension diagnosis/treatment:

- Have you ever been told by a doctor, nurse or health care professional that you have High blood pressure? (Yes/No/Refused/Don't know)
- We were previously told that you have High blood pressure, is this correct?
- Do you still have High blood pressure?
- Are you currently taking medication for High blood pressure? (Yes/No/Refused/Don't know)

- Definition of urban vs. rural household:

- Urban = Urban
- Rural = Traditional/Farms

| | | |
|---|---|---|
| 1 | Traditional | Communally owned land under the jurisdiction of traditional leaders. Settlements within these areas are villages. |
| 2 | Urban | A continuously built-up area that is established through township establishment such as cities, towns, 'townships', small towns, and hamlets. The areas are identified by "erf/erven/cadastre" from the Surveyor General or Municipal planning units. |
| 3 | Farms | Land allocated for and used for commercial farming including the structures and infrastructure on it. The areas are identified by farm and farm portion cadastre from the Surveyor General. |

### Appendix III: Classification of national education systems

| Survey name | None | Primary | Secondary | Tertiary |
|---|---|---|---|---|
| China Health and Nutrition Study (CHNS) | • Unschooled | • Elementary school | • Lower middle school<br>• Upper middle school | • Technical/Vocational school<br>• University/college degree<br>• Master's degree or higher |
| Indonesian Family Life Survey (IFLS) | • None<br>• Kindergarten | • Elementary school<br>• Adult education A<br>• Islamic Elementary school | • Junior high - general<br>• Junior high - vocational<br>• Senior high - general<br>• Senior high - vocational<br>• Adult education B & C<br>• Islamic Junior high school<br>• Islamic Senior high school | • Open University<br>• College D1, D2, D3<br>• University S1<br>• University S2<br>• University S3 |
| Mexican Family Life Study (MXFLS) | • No formal schooling<br>• Preschool or kindergarten | • Elementary | • Junior High<br>• Open Junior High School<br>• High School<br>• Open High School | • Trade School<br>• College<br>• Graduate |
| South African National Income Dynamics Study (NIDS) | • No schooling<br>• Grade 0/R | • Grades 1-7 | • Grade 8-12<br>• NTC 1<br>• NTC 2<br>• NTC 3<br>• Certificate with Grade 12 or less<br>• Diploma with less than Grade 12<br>• Certificate with Grade 12 | • Bachelor's degree<br>• Bachelor's degree and Diploma<br>• Honours degree<br>• Higher degree (Masters/Doctorate) |

### Appendix IV: Comparison of sample composition after exclusion of missing data

#### Appendix Table 1 Comparison of the samples after excluding those with missing data for China

| | **Age-eligible N=7380** | **Non-missing baseline information (after removing normotensive individuals and those missing baseline information) N = 2363** | **Analytic sample (additionally removing those who died, were lost to follow-up, or missing endline information) N=1371** |
|---|---|---|---|
| Age, mean (SD) | 56·9 (11·4) | 61·0 (11·3) | 59·1 (10·0) |
| Female, % (n) | 52 % (3874) | 51 % (1208) | 52 % (709) |
| Urban residence, % (n) | 35 % (2552) | 35 % (834) | 30 % (413) |
| Schooling, % (n) | | | |
| *None* | 29 % (2170) | 35 % (820) | 32 % (439) |
| *Primary* | 21 % (1533) | 22 % (512) | 24 % (334) |
| *Secondary* | 40 % (2949) | 35 % (830) | 36 % (493) |
| *Tertiary* | 9 % (678) | 8 % (201) | 8 % (105) |

**Appendix Table 2 Comparison of the samples after excluding those with missing data for Indonesia**

| | **Age-eligible N=14304** | **Non-missing baseline information (after removing normotensive individuals and those missing baseline information) N=5031** | **Analytic sample (additionally removing those who died, were lost to follow-up, or missing endline information) N=3438** |
|---|---|---|---|
| Age, mean (SD) | 55·1 (11·8) | 56·5 (11·4) | 54·1 (10·1) |
| Female, % (n) | 53 % (7632) | 58% (2899) | 60 % (2066) |
| Urban residence, % (n) | 52 % (7508) | 53% (2646) | 50 % (1724) |
| Schooling, % (n) | | | |
| *None* | 18 % (2549) | 19 % (979) | 17 % (598) |
| *Primary* | 49 % (7023) | 51 % (2555) | 53 % (1824) |
| *Secondary* | 25 % (3545) | 23% (1160) | 22 % (769) |
| *Tertiary* | 8 % (1088) | 7 % (337) | 7 % (247) |

### Appendix Table 3 Comparison of the samples after excluding those with missing data for Mexico

| | **Age-eligible N=10096** | **Non-missing baseline information (after removing normotensive individuals and those missing baseline information) N=2662** | **Analytic sample (additionally removing those who died, were lost to follow-up, or missing endline information) N=1946** |
|---|---|---|---|
| Age, mean (SD) | 55·9 (12·3) | 58·3 (11·9) | 57·4 (11·3) |
| Female, % (n) | 53 % (5342) | 59% (1570) | 62 % (1202) |
| Urban residence, % (n) | 58 % (5888) | 57% (1521) | 55 % (1072) |
| Schooling, % (n) | | | |
| *None* | 20 % (2072) | 22 % (598) | 21 % (416) |
| *Primary* | 52 % (5268) | 57 % (1511) | 59 % (1151) |
| *Secondary* | 18 % (1785) | 15 % (396) | 14 % (281) |
| *Tertiary* | 8 % (851) | 6 % (157) | 5 % (98) |

**Appendix Table 4 Comparison of the samples after excluding those with missing data for South Africa**

| | **Age-eligible N=7606** | **Non-missing baseline information (after removing normotensive individuals and those missing baseline information) N=3459** | **Analytic sample (additionally removing those who died, were lost to follow-up, or missing endline information) N=1604** |
|---|---|---|---|
| Age, mean (SD) | 55·7 (11·9) | 57·7 (11·7) | 54·8 (10·1) |
| Female, % (n) | 60 % (4560) | 67% (2323) | 73 % (1175) |
| Urban residence, % (n) | 52 % (3949) | 49% (1686) | 44 % (714) |
| Schooling, % (n) | | | |
| *None* | 27 % (2055) | 31 % (1061) | 29 % (458) |
| *Primary* | 34 % (2597) | 37 % (1280) | 39 % (633) |
| *Secondary* | 35 % (2677) | 30 % (1056) | 30 % (490) |
| *Tertiary* | 3 % (213) | 2 % (62) | 1 % (23) |

**Appendix V: Heatmap of transition probabilities from baseline to endline for each country**

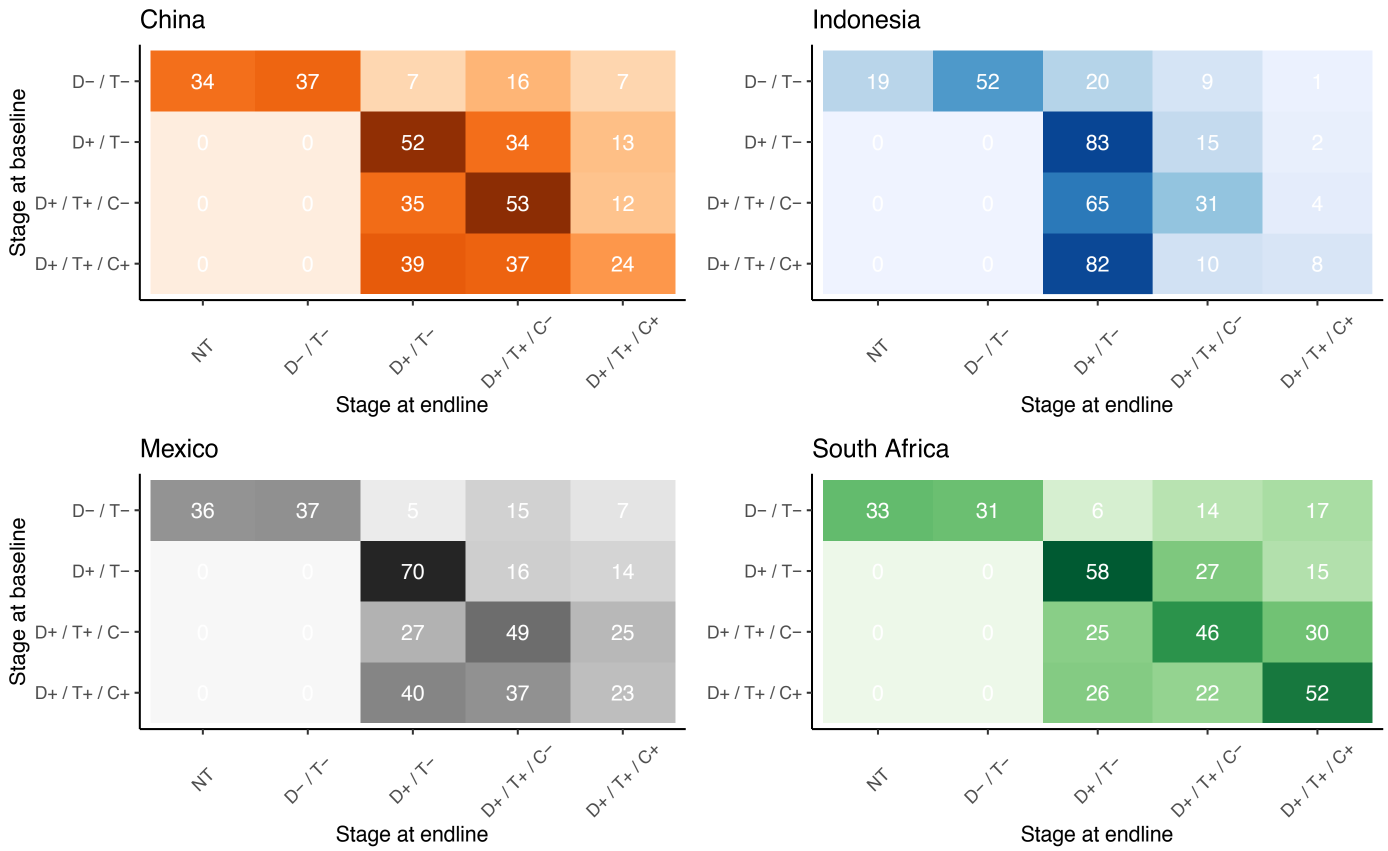


**Probability of transition between continuum stages from baseline to endline**

[a]Legend of continuum stages: NT = Normotensive, D-/T- = undiagnosed/untreated, D+/T- = diagnosed/untreated, D+/T+/C- = diagnosed/treated/uncontrolled, D+/T+/C+ = diagnosed/treated/controlled.

[b]Survey years: China (2009 to 2015), Indonesia (2007 to 2014), Mexico (2005/6 to 2009/12), South Africa (2008 to 2017).

[c]We weighted estimates in Indonesia, Mexico, and South Africa to be nationally representative; the China Health and Nutrition Survey does not contain survey weights.

**Appendix VI:  Successive stages of the hypertension care continuum**

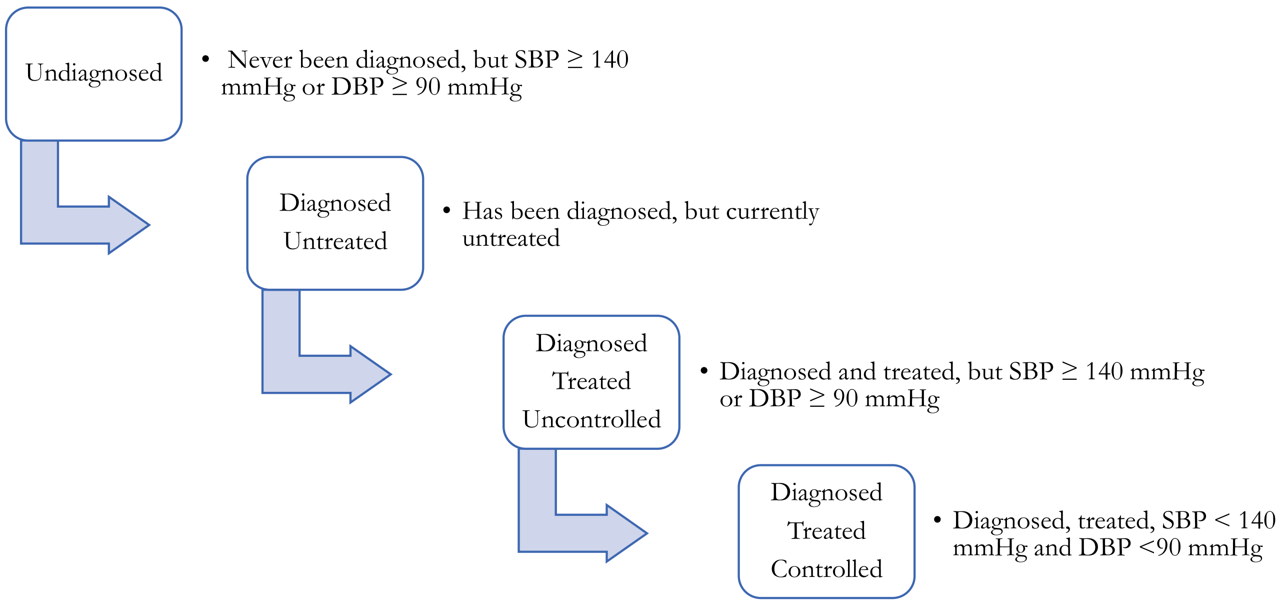
